# Genetic analysis of 42 Y-STR loci in Han and Manchu populations from the three northeastern provinces in China

**DOI:** 10.1101/2023.06.19.23291583

**Authors:** Wenqian Song, Shihang Zhou, Weijian Yu, Yaxin Fan, Xiaohua Liang

## Abstract

**Background:** Y-STR polymorphisms are useful in tracing genealogy and understanding human origins and migration history. This study aimed to fill a knowledge gap in the genetic diversity, structure, and haplogroup distribution of the Han and Manchu populations from the three northeastern provinces in China (Liaoning, Jilin, and Heilongjiang).

**Methods:** A total of 1,048 blood samples were collected from unrelated males residing in Dalian. Genotyping was performed using the AGCU Y37+5 Amplification Kit, and the genotype data were analyzed to determine allele and haplotype frequencies, genetic and haplotype diversity, discrimination capacity, and haplotype match probability. Population pairwise genetic distances (*F*_*st*_) were calculated to compare the genetic relationships among Han and Manchu populations from Northeast China and other 36 populations using 27 Yfiler Plus loci set. Multi-dimensional scaling and phylogenetic analysis were employed to visualize the genetic relationships among the 40 populations. Moreover, haplogroups were predicted based on 27 Yfiler Plus loci set.

**Results:** The Han populations from Northeast China exhibited genetic affinities with both Han populations from the Central Plain and the Sichuan Qiang population, despite considerable geographical distances. Conversely, the Manchu population displayed a relatively large genetic distance from other populations. The haplogroup analysis revealed the prevalence of haplogroups E1b1b, O2, O3, and Q in the studied populations, with variations observed among different ethnic groups.

**Conclusion:** The study contributes to our understanding of genetic diversity and history of the Han and Manchu populations in Northeast China, the genetic relationships between populations, and the intricate processes of migration, intermarriage, and cultural integration that have shaped the region’s genetic landscape.

## Background

Northeast China usually refers to the three northeastern provinces in China including Liaoning, Jilin, and Heilongjiang. As one of the major port cities of Northeast China, Dalian experienced an extreme population loss during World War I. Therefore, most of Dalian’s current inhabitants are immigrants from other areas. Over recent decades, Dalian has attracted a considerable population from the three northeastern provinces due to its advanced economic development and plentiful job opportunities (Zuo G. 2020) (Fig 1). At present, the Han ethnic group constitutes the largest segment of Dalian’s population at 84%, followed by the Manchu group at 13% (Adnan et al. 2020).

**Fig 1.**
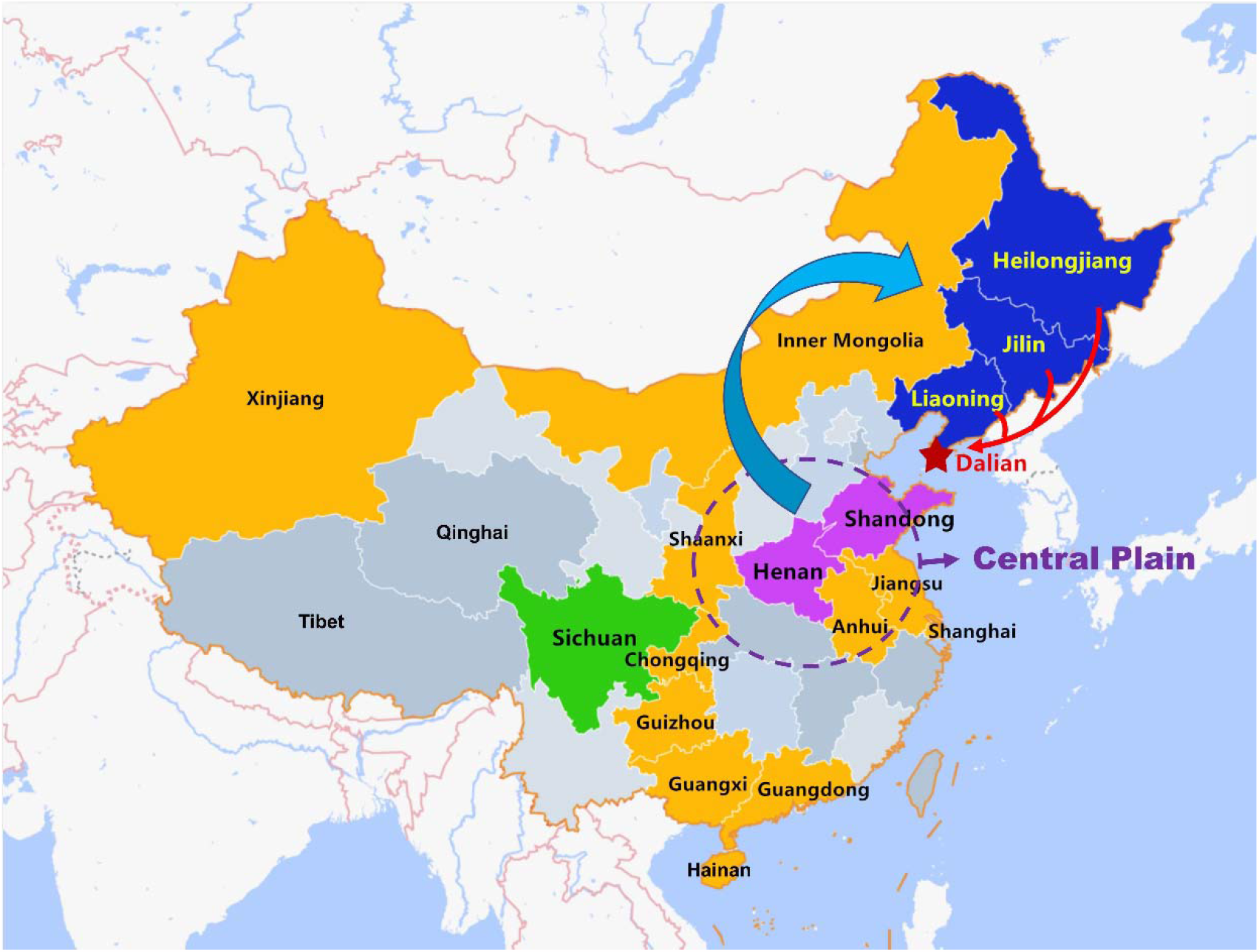
Geographical locations of populations from Northeast China and other Chinese populations included in this study. The cyan and red arrows indicate the migration of Han populations. Dalian, marked as a red pentagram, is located in geographic coordinates 38°43′∼40°12′n, 120°58′∼123°31′e

The Central Plain, often regarded as the cradle of Chinese civilization, typically encompasses regions surrounding the Yellow River, including areas such as Henan, Shandong, and Hebei (Cioffi-Revilla & Lai 1995; Duara 2003). Historically, the Manchu people, originating from Northeast China, became the ruling class across all of China during the Qing dynasty (1636-1912 AD). A segment of the Manchus then transitioned from Northeast China to the Central Plain, as the government aimed to solidify its control over the country. This shift resulted in the Manchus’ partial assimilation into the Han Chinese population (Elegant 2017).

Before the Qing dynasty, the Han Chinese were a small minority in Northeast China. However, in the late 19th century, the Qing government encouraged the development of Northeast China to prevent the Russian invasion, which led to a massive influx of people from the Central Plain to Northeast China for seeking new economic opportunities (Tian F. & Chen Y. 1986) (Fig S1). As a result of these substantial migrations during the late Qing dynasties and the early Republic of China (1860-1931), the Han became the dominant ethnic group in Northeast China. Over several centuries, more than 20 million people have migrated to Northeast China, with the vast majority (up to 90%) coming from Shandong and Henan provinces. Consequently, the majority of the current Han inhabitants of Northeast China can trace their roots back to the Central Plains (Gao L. 2005).

Today, about 97% of the Manchu population resides in Northeast China, Inner Mongolia, and Beijing, with the largest concentration in Liaoning Province, the birthplace of the Manchu people (Liu 1995). Despite this, the Manchu language is on the verge of extinction, as many regional Manchus have lost the ability to speak it (He & Guo 2013).

Y-chromosomal short tandem repeats (Y-STRs) are widely employed in forensic DNA analysis, serving as a valuable tool in defining paternal lineages in situations such as identifying male suspects in sexual assault cases (Kayser 2017). Additionally, Y-STR polymorphisms are useful in tracing genealogy and understanding human origins and migration history (Underhill & Kivisild 2007). Haplogroups are branches on the tree of early human migrations and genetic evolution. They cluster individuals who share a common ancestor and possess specific genetic mutations, which contributes to our understanding of human migration and evolution (Kapur et al. 2006).

This study aimed to fill a knowledge gap in the genetic diversity, structure, and haplogroup distribution of the Han and Manchu populations in Northeast China. We genotyped 1048 males residing in Dalian city using a 42 Y-STR system and investigated the genetic diversity and structure of this population. Among them, the genetic relationships of Han and Manchu populations from Northeast China were compared with various ethnic groups in different regions and countries based on the 27 Y-STR system. Next, we explored the Y-STR haplogroup distribution of Northeast Han and Man populations and several other populations throughout China.

## Results and Discussion

### Forensic characteristics

A total of 1048 unrelated males in Dalian were genotyped using AGCU Y37+5 Amplification Kit, which includes 27 Yfiler Plus loci. In order to estimate the genetic diversity for Y-STRs in 958 Han individuals (818 from Northeast China, 140 from other regions) and 68 Manchu individuals in Dalian (Table S1 and S2), allele frequency and forensic parameters including GD and DC were calculated for each locus incorporated in the AGCU Y37+5 panel (Table S3 and S4). For both Han and Man populations in Dalian, the GD of all the loci was above 0.4 except DYS645 and 5 indel loci (Fig 2). Three multi-locus Y-STRs including DYS527, DYS385, and DYF387S1 showed the highest GD values of more than 0.9.

**Fig 2.**
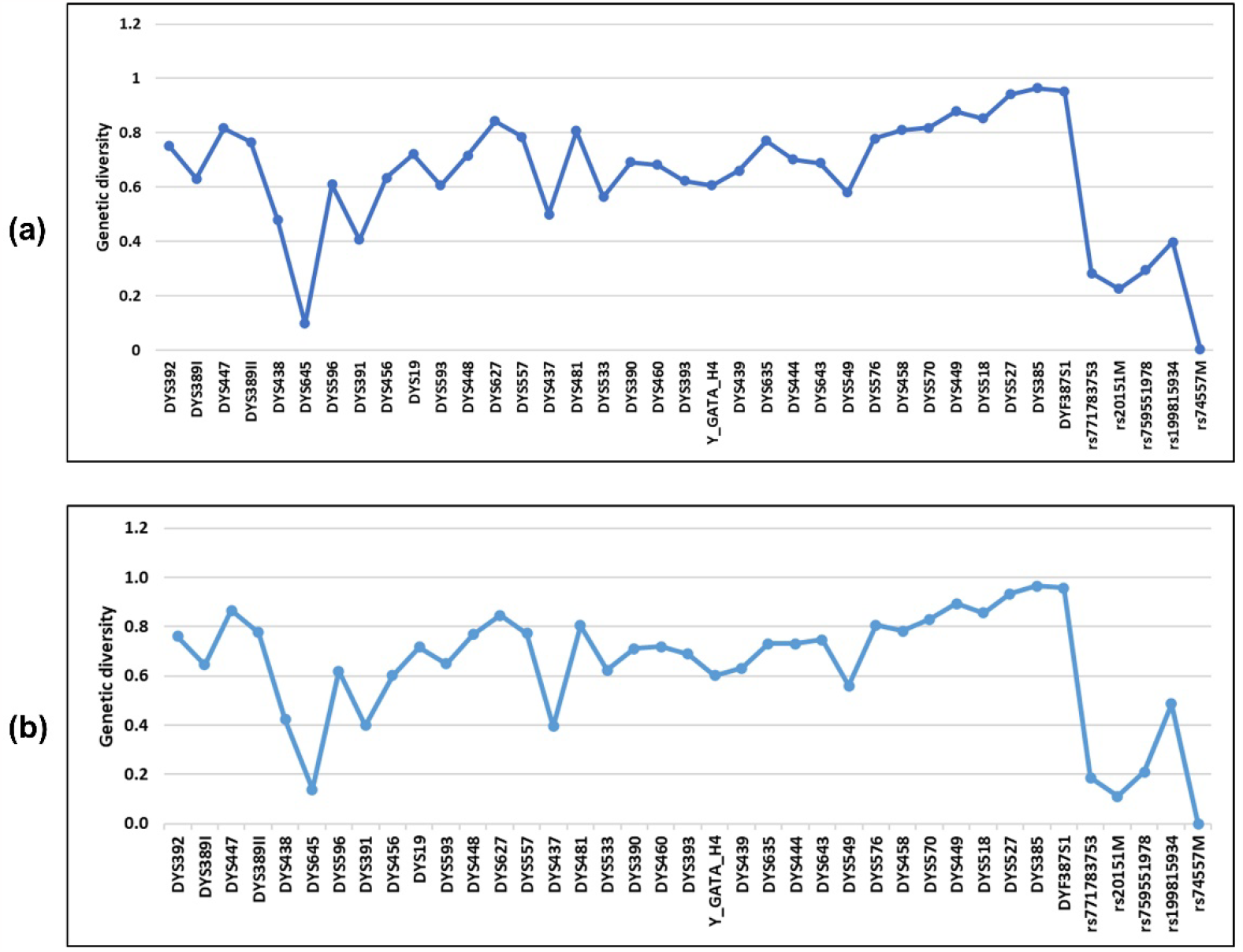
Genetic diversity (GD) across 40 loci. (a) Han population in Dalian (n=958); (b) Man population in Dalian (n=68)

To assess the Y-STR haplotype DC, HD, and HMP of 958 Han individuals in Dalian, haplotype information was analyzed in different combinations of Y-STR loci, including minimal nine loci, PowerPlex Y12 loci, 17 Yfiler loci, PowerPlex Y23 loci, 27 Yfiler Plus loci and 29 Ymax loci (Table S5). As more Y-STR loci were included in the analysis, the percentage of unique haplotypes, HD, and DC increased, which means that the individuals had more diverse haplotypes. At the same time, HMP decreased, indicating a reduced likelihood of two individuals having matching haplotypes. Therefore, incorporating an increased number of Y-STR loci in genetic analysis enhances the precision and dependability of discerning genetic associations among individuals. However, the 27 Yfiler Plus loci set and the 29 Ymax loci set were shown to yield identical haplotype numbers, GD, HD, and HMP values, corroborating findings from previous studies (Jung et al. 2016; Zhou et al. 2018). This indicates that beyond a certain threshold, the addition of extra loci may not significantly augment the accuracy of genetic examinations, especially when dealing with finite sample size.

### Genetic structure and population comparisons

This population-specific analysis was focused on specific groups of unrelated male individuals born in Northeast China. These groups include the Han population from Liaoning (n = 571), Jilin (n = 89), Heilongjiang (n = 158), and the Manchu population (n = 68) (Table S1 and S2). Pairwise genetic distances (*F*_*st*_) and corresponding p-values were calculated among these four groups and the other 36 populations, utilizing a 27 Yfiler Plus loci set (Table S6 and S7). Out of the 40 populations included in this study, the Han population from Jilin and Shandong have the least genetic distance (*F*_*st*_ = 0.00080), whereas the Basque population from Europe and the Arabian population from South Asia exhibit the largest genetic distance (*F*_*st*_ = 0.27972). The heatmap provides a visual representation of the degree of genetic differentiation between populations (Fig 3). From this, it is apparent that the Kyrgyz population from Xinjiang, the Arabian population from South Asia, and the Basque population from Europe show the most significant genetic differences compared to the other 37 populations.

**Fig 3.**
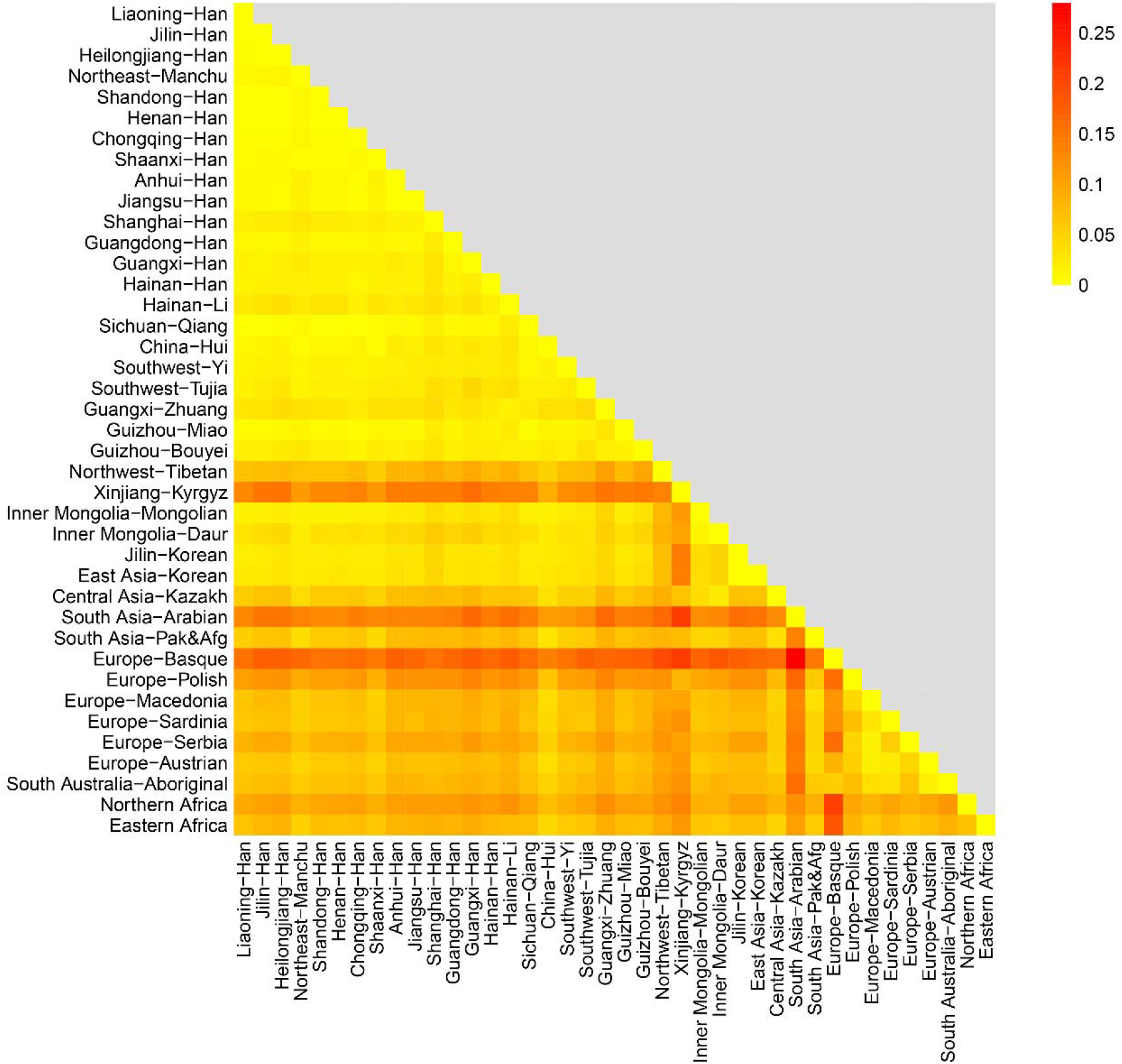
Genetic differentiation degree between 40 populations based on analysis of 27 Yfiler Plus loci

Among the Han population from the three northeast provinces in China, the Jilin subgroup exhibited a relatively smaller genetic distance with Liaoning (*F*_*st*_ = 0.00167) and Heilongjiang subgroups (*F*_*st*_ = 0.00119). Conversely, the genetic distance between Liaoning and Heilongjiang subgroups was comparatively larger (*F*_*st*_ = 0.00306). Within the broader scope of China, the Northeast Han populations show the highest genetic affinity with Shandong Han (0.00080 ≤ *F*_*st*_ ≤ 0.00113), Henan Han (0.00137 ≤ *F*_*st*_ ≤ 0.00266), and Sichuan Qiang (0.00188 ≤ *F*_*st*_ ≤ 0.00414) populations. In particular, the Shandong Han population displays a closer genetic relationship to the Han populations from Jilin (*F*_*st*_ = 0.00080) and Heilongjiang (*F*_*st*_ = 0.00082) than to those from Liaoning (*F*_*st*_ = 0.00113). Conversely, the Henan Han population exhibits a more pronounced genetic similarity to the Liaoning Han population (*F*_*st*_ = 0.00137), as compared to the Jilin Han (*F*_*st*_ = 0.00236) and the Heilongjiang Han (*F*_*st*_ = 0.00266) populations. These findings suggest that during the migrations of the Han populations from the Central Plain to Northeast China, a larger number of individuals from Shandong chose to relocate to Jilin and Heilongjiang, whereas a considerable portion of individuals from Henan decided to settle in Liaoning.

Additionally, the Sichuan Qiang population demonstrates a genetic affinity with the Han populations from Liaoning (*F*_*st*_ = 0.00199), Jilin (*F*_*st*_ = 0.00188), Shandong (*F*_*st*_ = 0.00214), Henan (*F*_*st*_ = 0.00263), and Chongqing (*F*_*st*_ = 0.00150). Notably, Sichuan is geographically distant from all these regions, except for Chongqing. Although several studies have uncovered evidence of a close genetic relationship between the Qiang and Han groups, none have explored this genetic relationship considering potential gene flow and geographic location (Song et al. 2022; C.-C. Wang et al. 2014; Wu et al. 2020). In this study, we found that the Sichuan Qiang population displays closer genetic proximity to the Han population from Northeast China (Liaoning and Jilin) than to those from the Central Plains (Shandong and Henan), despite the Northeast Han population’s migration from the Central Plains. Such a pattern suggests the possibility of substantial gene flow or migration between these populations in the past. However, the underlying cause of this paticular genetic proximity remains uncertain due to the absence of corresponding historical records.

According to historical records, the Qiang people are a minority ethnic group in China who traditionally lived in the mountainous regions of western China, including the areas around the Min River in Sichuan province for thousands of years (Wang 2013). In the 17th century, Han people began to move into the Qiang’s territory, primarily for economic reasons such as farming and trade. This led to increased interaction between the Han and Qiang people, and over time, intermarriage between the two groups became more common. As a result, numerous Qiang people today have some Han ancestry (Wen 2014). Intriguingly, the ethnic identity of the Han Chinese population in China is typically primarily inherited through the patrilineal lineage, which is reflected in the Y-chromosome DNA. However, based on the result of this study, it appears that many Han males who intermarry with Qiang individuals or their descendants tend to adopt Qiang cultural identity over their own Han cultural identity. Accordingly, in the process of multi-ethnic integration, people’s identification with ethnicity is not strictly based on paternal lineage, whereas it is more of a sociological concept than a matter of racial bloodline.

Furthermore, our study includes a Manchu population, about 91% of which is from Liaoning. Even though Han and Manchu people cohabit in Liaoning, they display a relatively substantial genetic difference (*F*_*st*_ = 0.00575). The genetic distance between the Manchu and Han populations increases as one moves northwards from Liaoning towards Heilongjiang. The Manchu population presents a relatively large genetic distance from all the other 39 populations examined in the study (*F*_*st*_ > 0.00500). Theoretically, the commencement of mutual integration between the Han and Manchu groups should have taken place in the 1600s during the Qing dynasty, when the Manchu people ascended to the ruling class of China. However, to maintain the purity and nobility of their ethnic bloodline as well as preserve their traditional language and customs, the Manchu rulers implemented a policy that barred intermarriage between the Manchu and Han peoples (Jian 2017). This policy, in effect, impeded the integration process between these two ethnic groups.

Pairwise genetic distances between 40 populations estimated by 27 Yfiler Plus set were visualized in the MDS plot (Fig 4). The MDS plot reinforces the genetic affinity between the Han populations from the Northeast China and Central Plain, and the Sichuan Qiang population. These groups also demonstrate genetic proximity to the Guizhou Miao population as well as the Han populations from Chongqing, Jiangsu, Anhui, Guangdong, Hainan, and Shaanxi. Conversely, despite their geographical location within China’s territory, the Northwest Tibetan, Inner Mongolia Daur, and Xinjiang Kyrgyz populations appear genetically distinct, indicating their isolation from the aforementioned cluster.

**Fig 4.**
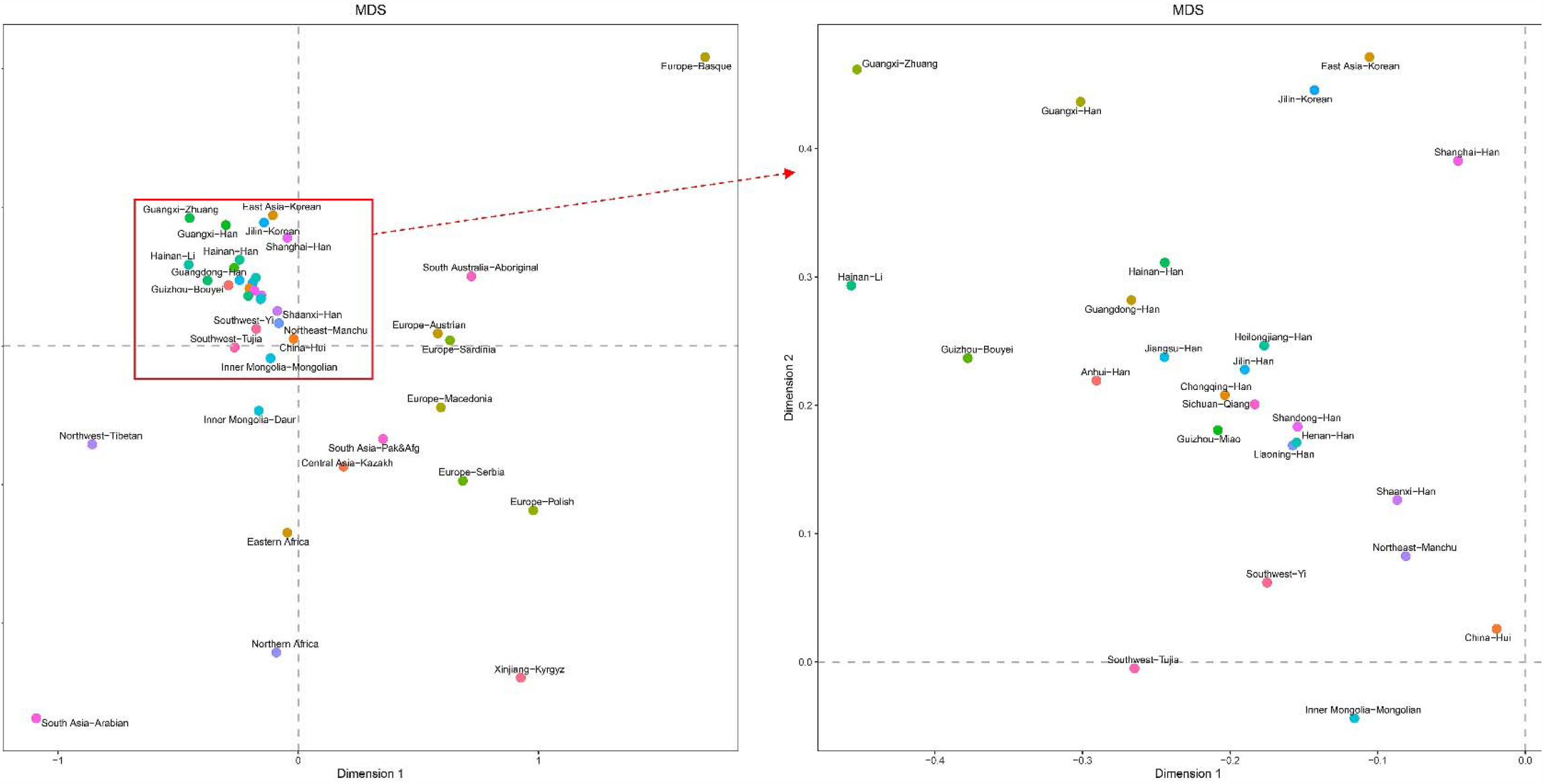
MDS plot between 40 populations based on pairwise Fst values using 27 Yfiler Plus set

Subsequently, we constructed an N-J phylogenetic tree based on the *F*_*st*_ values to further verify the genetic relationships among the 40 populations (Fig 5). Generally, the phylogenetic structures are in line with the MDS analysis outcomes. The majority of the Chinese populations formed a single large cluster, except the Northwest Tibetan and Xinjiang Kyrgyz populations. Several geographically proximate populations formed tight clusters on the tree, including Sichuan Qiang and Chongqing Han, Henan Han and Shandong Han, Heilongjiang Han, and Jilin Han, as well as Jiangsu Han and Anhui Han. Interestingly, the Liaoning Han population initially clustered with the Henan Han and Shandong Han populations, then with the Chongqing Han, Sichuan Qiang, and Guizhou Miao populations, and finally with the Jilin Han and Heilongjiang Han populations. This pattern suggests a certain degree of phylogenetic isolation of the Liaoning Han from the Jilin and Heilongjiang Han populations. This divergence may be attributed to a higher level of population movement in Liaoning, potentially driven by favorable economic opportunities. It is worth noting that the Northeast Manchu and Shaanxi Han populations form a tight cluster, despite a relatively large genetic distance (*F*_*st*_ = 0.00534). This may be attributed to specific historical intermarriage or migration patterns. Nevertheless, the small sample size of the Northeast Manchu population may lead to misleading results.

**Fig 5.**
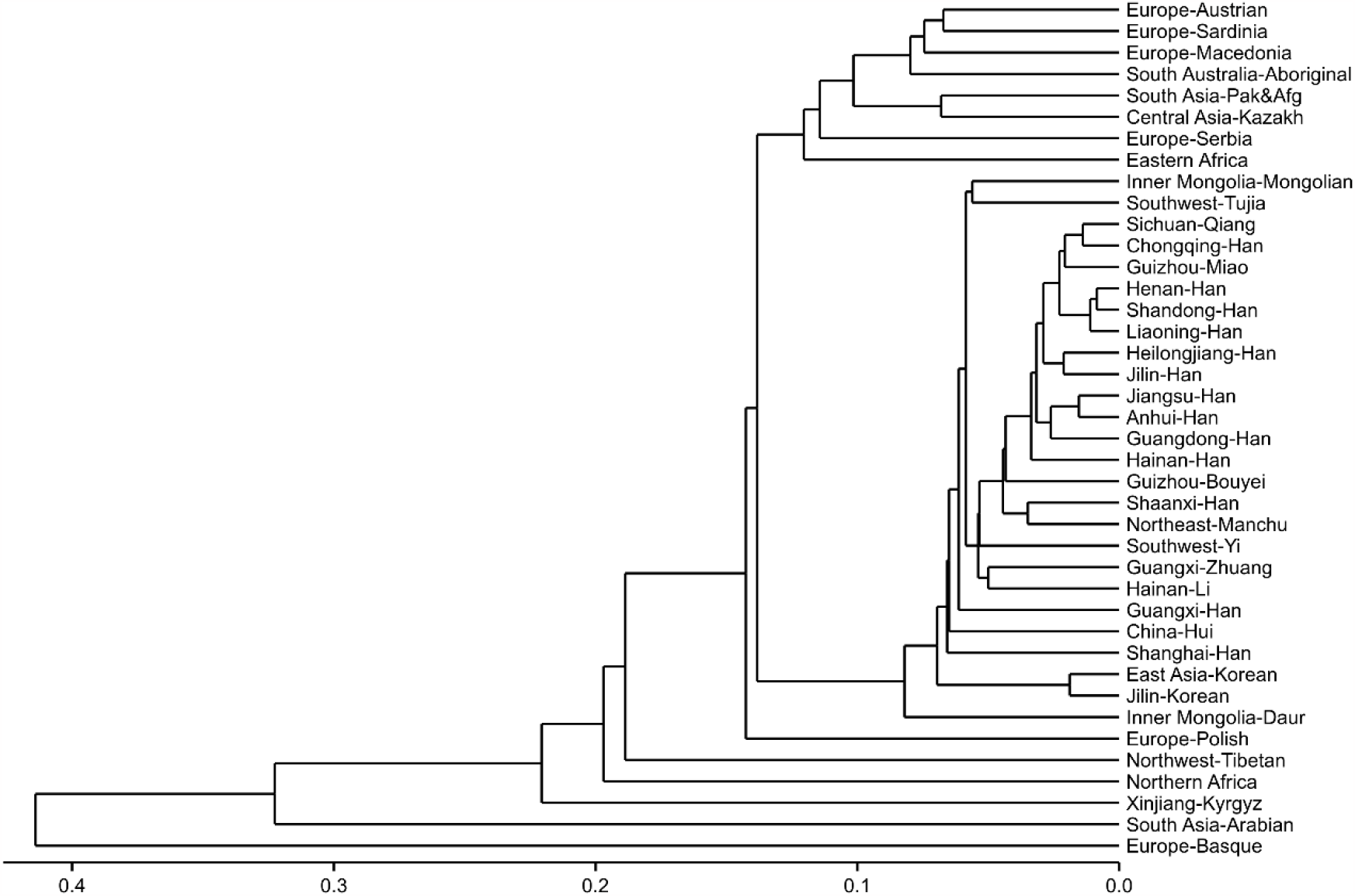
The neighbor-joining phylogenetic tree of 40 populations based on pairwise Fst values using 27 Yfiler Plus set

### Y-chromosome haplogroup prediction

A total of 818 Northeast Han (Liaoning Han, n = 571; Jilin Han, n = 89; Heilongjiang Han, n = 158) and 68 Manchu individuals as well as 13 other Chinese populations were assigned to haplogroups based on 27 Yfiler Plus loci using the Whit-Athey algorithm with the Haplogroup Predictor tool. A total of 27 Asian haplogroups were identified, including C3, E1a, E1b1a, E1b1b, G1, G2a, G2c, H, I1, I2a (xI2a1), I2a1, I2b (xI2b1), I2b1, J1, J2a4b, J2a4h, J2a4 (x bh), J2b, L, N, O2, O3, Q, R1a, R1b, R2 and T (Table S8). The haplogroup distribution of 17 populations was visualized with varying ethnicities and geographical locations within China (Fig 6). The major haplogroups identified in the study were E1b1b (30%-35%), O2 (21%-39%), O3 (8%-11%), and Q (9%-16%) for the populations from Northeast China, which accounted for more than 75% of the Y lineages. It appears that the two haplogroups G2c and J2a4h were absent in all 17 populations.

**Fig 6.**
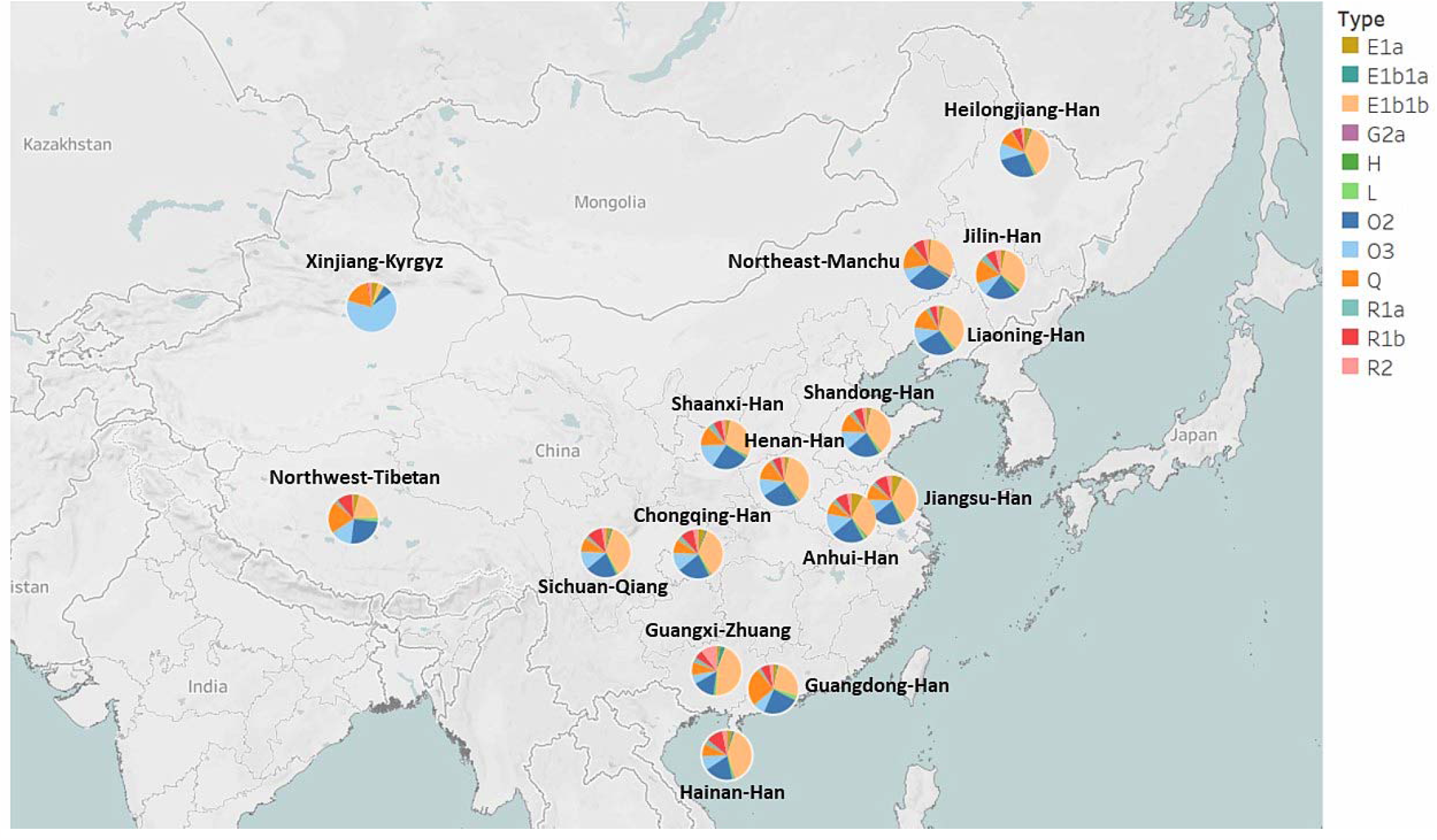
Haplogroup distribution in the 17 geographical populations. Haplogroups with a fitness probability less than 1% are not included in the figure. Different colors represent the fitness probability of each haplogroup

In this study, the predominant haplogroup in the populations, except for the Tibetan and Kyrgyz groups, is E1b1a. Haplogroup E1b1b traces its origin back to East Africa before becoming widespread in Eurasia (Bajić et al. 2018). We found that Guangxi Zhuang and Hainan Han populations notably display a higher percentage of E1b1b, at 39% and 44% respectively, in contrast to the Guangdong Han population, which has a lower percentage of E1b1b at 26%. Among the Northwest Tibetan and Xinjiang Kyrgyz populations, the presence of E1b1a is less common, with percentages of 20% and 4%, respectively. The next most prevalent haplogroups in all Chinese populations, except for the Kyrgyz group, are O2, O3, or Q.

Our findings slightly differ from previous studies, which have indicated that haplogroups O2 and O3 are the most prevalent haplogroups among East Asians (Deng et al. 2004; Fan et al. 2023; Y. Wang et al. 2015). This discrepancy may result from the use of different marker combinations or haplogroup prediction models. Deng et al. (2004) investigated Chinese Y haplogroups using nested cladistic analysis based on non-recombining portions of the Y-chromosome (NRY) SNPs, while Fan et al. (2023) employed a knearest neighbor predictor based on 23 common Y-STR loci. Wang et al. (2015) utilized Whit Athey’s Haplogroup Predictor based on 17 Y-STRs. In contrast, our study used 27 Yfiler Plus loci as markers submitted to Whit Athey’s Haplogroup Predictor.

Additionally, in our study, the Kyrgyz population displays a higher prevalence of haplogroup O3 (64%) and a significantly lower percentage of E1b1a (4%) compared to other Chinese populations. This observation implies that the Kyrgyz people may have a unique genetic history that sets them apart from other populations in China. Furthermore, six haplogroups (I1, I2a1, J2a4b, J2a4 (x bh), J2b, and N) were not observed in the Manchu population, in contrast to other Chinese populations. This may be indicative of limited intermarriage between the Manchu population and other groups. However, it is important to consider that the small sample size of the Manchu population could lead to less precise estimates of the population’s haplogroup diversity.

## Conclusion

In summary, this study investigated the genetic diversity, structure, and haplogroup distribution of the Han and Manchu populations from Northeast China. The results revealed several key findings. First, the Han populations from Northeast China exhibited genetic affinities with Han populations from the Central Plain and Qiang population from Sichuan, indicating historical migrations, substantial gene flow, and intermarriage among these groups. Second, the Manchu population displayed distinct genetic characteristics and maintained a relatively large genetic distance from other populations, possibly due to historical policies and limited intermarriage with the Han population. Finally, the haplogroup distribution analysis revealed the prevalence of haplogroups E1b1b, O2, O3, and Q in the studied populations, with variations observed among different ethnic groups. Overall, this research contributes to our understanding of the genetic diversity and history of the Han and Manchu populations from Northeast China. The findings shed light on the genetic relationships among these populations and provide insights into the complex processes of migration, intermarriage, and cultural integration that have shaped the genetic landscape of the region.

## Methods

### Sample preparation

A total of 1048 blood samples from unrelated Chinese males were collected from Dalian, Liaoning Province, China. Their ethnic groups and birthplaces were confirmed through their ID cards and self-declarations (Liaoning Han, n = 571; Jilin Han, n = 89; Heilongjiang Han, n = 158; Northeast China Manchu, n = 68; Others, n = 162). All participants signed informed consent. All the experimental procedures were performed following the standards of the Declaration of Helsinki. This study was approved by the Medical Ethics Committee of Dalian Blood Center.

### Y-STR multiplex genotyping

Human genomic DNA was directly amplified from FTA cards (Whatman Inc., Clifton, NJ, US) by AGCU Y37+5 Amplification Kit (AGCU, Wuxi, China) according to the manufacturer’s instructions using TC-96 PCR Thermal Cycler (Bioer Technology, Hangzhou, China). The AGCU Y37+5 Amplification Kit includes 27 Yfiler Plus loci (DYS576, DYS389I, DYS635, DYS389II, DYS627, DYS460, DYS458, DYS19, YGATAH4, DYS448, DYS391, DYS456, DYS390, DYS438, DYS392, DYS518, DYS570, DYS437, DYS385a/b, DYS449, DYS393, DYS439, DYS481, DYF387S1a/b, DYS533) plus 10 highly polymorphic Y-STR loci (DYS444, DYS447, DYS527a/b, DYS557, DYS576, DYS593, DYS596, DYS643, DYS645) and 5 Y-indel loci (rs199815934, rs20151M, rs74557M, rs759551978, rs771783753). The amplified products were detected by capillary electrophoresis on the ABI-3730 Genetic Analyzer (Thermo Fisher Scientific, Waltham, MA, US). The data were analyzed by the GeneMapper ID -X (Thermo Fisher Scientific, Waltham, MA, US). The control DNA 9948 was genotyped for quality control purposes.

### Statistical analyses

Allele and haplotype frequencies, genetic diversity (GD), and haplotype diversity (HD) were computed by GenAlEx 6.5 software. Discrimination capacity (DC) and haplotype match probability (HMP) were calculated following the given formulae (Table S3, S4, and S5). Based on 27 Yfiler Plus loci, population pairwise genetic distance (*F*_*st*_) and *p* values were estimated by analysis of molecular variance (AMOVA) using Arlequin v3.5 software. This allowed us to compare the genetic distances between the 4 populations under study and 36 other populations reported in the Y-Chromosome STR Haplotype Reference Database (YHRD). Then a heatmap was generated using R-4.2.2 program (R Core Team), where each cell is colored corresponding to the magnitude of the pairwise *F*_*st*_ values. Multi-dimensional scaling (MDS) was also employed to create a reduced dimensionality spatial representation of populations based on *F*_*st*_ values by R-4.2.2 program (R Core Team). Additionally, phylogenetic relationships among 40 populations were depicted by a neighbor-joining (N-J) phylogenetic tree using R-4.2.2 program (R Core Team). Furthermore, based on 27 Yfiler Plus loci, the haplogroups of 17 populations in China were identified with the 27-Haplogroup Program in Whit Athey’s Haplogroup Predictor (http://www.hprg.com/hapest5/). Then Tableau 2021.1 was employed to visualize the distribution of haplogroups across various populations.

## Data Availability

All data produced in the present study are available upon reasonable request to the authors

## Declarations

### Ethics Approval and Consent to Participate

This study was performed in line with the principles of the Declaration of Helsinki. Approval was granted by the Ethics Committee of Dalian Blood Center. Informed consent was obtained from all individual participants included in the study.

### Consent for Publication

Not applicable.

### Availability of data and materials

The datasets generated and analyzed during the current study are available from the corresponding author on reasonable request.

### Author Contributions

All authors contributed to the study conception and design. Material preparation, data collection and analysis were performed by Wenqian Song, Shihang Zhou, Yaxin Fan, Weijian Yu and Xiaohua Liang. The first draft of the manuscript was written by Wenqian Song and all authors commented on previous versions of the manuscript. All authors read and approved the final manuscript.

## Acknowledgments

We are grateful to all the volunteers for providing samples for this study.

